# Bioaersols in orthopedic surgical procedures and implications for clinical practice in the times of COVID-19 pandemic: a protocol for systematic review and meta-analysis

**DOI:** 10.1101/2020.07.19.20157511

**Authors:** Siddhartha Sharma, Rakesh John, Deepak Neradi, Sandeep Patel, Mandeep Singh Dhillon

## Abstract

**Background:** Orthopedic surgical procedures involve a number of aerosol generating procedures; these include electrocautery, power instruments for bone cutting, burring and drilling, and tools for wound lavage. This assumes a great significance in the context of the current COVD-19 pandemic, as there are chances of aerosol-borne disease transmission in orthopedic surgical procedures. Hence, this systematic review and meta-analysis will be undertaken to assimilate and analyse the available evidence on bioaerosols in orthopedic surgical procedures and their significance with respect to SARS-CoV-2 virus transmission.

**Objectives:** To determine the characteristics (amount and/or density, size, infectivity, and spread etc.) of bioaerosols found in orthopaedic operating rooms (ORs) and to determine the characteristics of aerosols generated by different orthopaedic power tools and devices.

**Methods:** A systematic review and meta-analysis will be conducted. The PRISMA guidelines will be strictly followed. The primary search will be conducted on the PubMed, EMBASE, Scopus, Cochrane Library, medRxiv, bioRxix and Lancet preprint databases, using a well-defined search strategy. Any original research study (including cohort, case-control, case series, cadaveric studies and studies, animal models, laboratory based experimental studies) looking at aerosol generation in orthopedic surgical procedures, or aerosol generation by orthopaedic power tools and devices will included. Outcome measures will include characteristics (amount and/or density, size, infectivity, and spread etc.) of bioaerosols found in orthopaedic operating rooms (ORs) and those generated by various orthopaedics power tools and devices. Metanalysis using the random-effects model will be conducted to determined pooled estimates of the outcome variables. Heterogeneity will be assessed by the *I*^*2*^ test. Risk of bias will be assessed by the Risk of Bias in Studies estimating Prevalence of Exposure to Occupational risk factors (RoB-SPEO) tool. The overall strength of evidence will be assessed by the GRADE approach.

## 1. Background

The SARS-CoV-2 virus has caused an unprecedented, catastrophic pandemic, which has adversely affected healthcare systems globally (1). As a result, the number of surgical procedures in hospitals has decreased substantially, owing in part to the potential risk of viral transmission through aerosols (2,3). Orthopaedic surgical procedures (OSP) are considered as aerosol generating procedures (AGP), chiefly due to the use of high-speed drilling and cutting tools, electrocautery and wound lavage systems (4,5). However, there is a dearth of supporting scientific data about the quantity, quality and infectivity of the aerosols generated during OSP (4).

## 2. Need for review

The proposed systematic review and meta-analysis will be conducted to assimilate and summarize the existing literature on aerosol generation in orthopedic procedures. In the context of the current COVID-19 pandemic, we also wish to analyse the available evidence to determine what preventive strategies could be undertaken to decrease the risk of transmission of the SARS-CoV-2 virus through bioaerosols generated during orthopedic surgical procedures.

## 3. Objectives

This study has two key objectives:

a. To determine the characteristics (amount and/or density, size, infectivity, and spread etc.) of bioaerosols found in orthopaedic operating rooms (ORs).
b. To determine the characteristics of aerosols generated by different orthopaedics power tools and devices.

## 4. PICO framework for the study

a. Participants: patients/animals undergoing orthopedic surgical procedures (depending on study design)
b. Intervention: orthopaedics surgical procedures
c. Control: none
d. Outcomes: various aspects of aerosol generation, as outlined in section 5(f)

## 5. Methods

This systematic review and meta-analysis will be conducted in accordance with the Preferred Reporting Items for Systematic Reviews and Meta-analysis (PRISMA) guidelines.

a. *Review Protocol* A protocol of the systematic review will be formulated a priori and made available online. The review protocol will be formulated in accordance with the PRISMA-P guidelines. **(Appendix I)**
b. *Eligibility Criteria* Any original research study (including cohort, case-control, case series, cadaveric studies and studies, animal models, laboratory based experimental studies) looking at aerosol generation in OSP or aerosol generation by orthopaedic power tools will included. Both English and non-English articles will be included. Studies looking at AGPs in specialties other than orthopaedic surgery, conference papers, review articles, expert or personal opinions and editorials will be excluded. Studies using which do not evaluate aerosol generation in orthopaedic surgical procedures, including ‘*sham-surgery*’ designs will also be excluded.
c. *Information Sources & Literature search* The primary search will be conducted on the PubMed, EMBASE, Scopus and Cochrane Library databases, using a well-defined search strategy. The medRxiv (https://www.medrxiv.org/), bioRxiv (https://www.biorxiv.org/) and Lancet preprint (https://www.thelancet.com/preprints) servers will also be searched to identify unpublished studies **(Table 1)**. For the secondary search, a manual search of references from the full-text of all included articles & relevant review articles will be conducted. There will be no restrictions on the language or date of publication. Furthermore, the electronic databases of select peer-reviewed orthopaedics journals (JBJS American, Bone and Joint Journal, Journal of Orthopaedic Trauma, Clinical Orthopaedics and Related Research, Injury and Acta Orthopaedica) will also be searched to identify relevant literature.

**Table 1:**
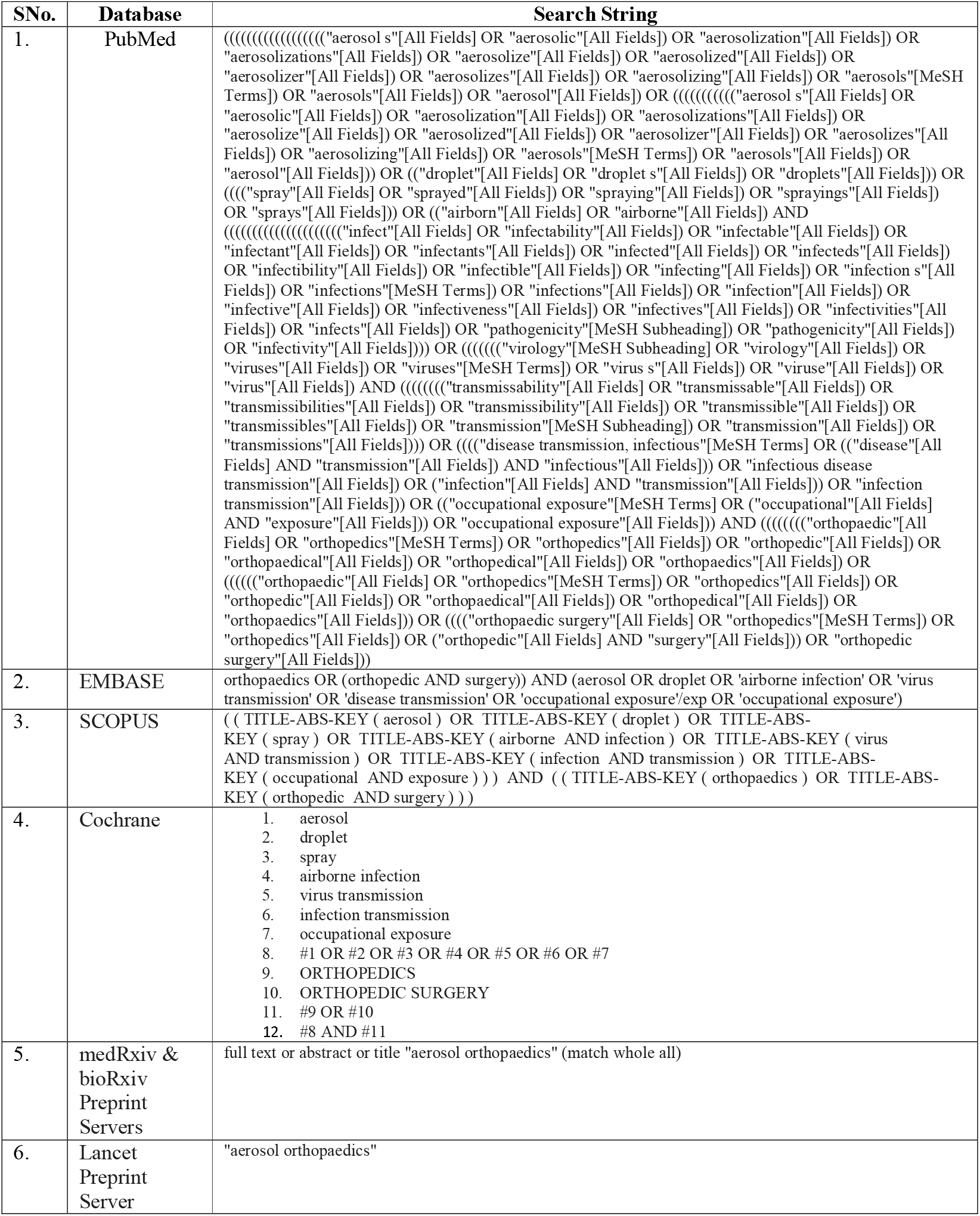
Literature search strategy for the review
d. *Study Selection* Two authors (SS and RJ) will independently screen titles and abstracts of all articles identified in the initial search. The full-texts of all articles shortlisted after initial screening will be obtained, assessed against the prespecified inclusion and exclusion criteria, and accepted or rejected, as appropriate. In the event of disagreement, a consensus will be reached by discussion with the intervention of the senior author (MSD). If needed, the author(s) of eligible studies will be contacted for clarification to determine inclusion or exclusion in the review. Reasons for exclusion of those studies for which full-text was obtained will be documented.
e. *Data Collection & Data Items* Two authors (SS and RJ) will extract data from each included study independently using pre-piloted data extraction forms; this will be cross-checked by a third author (DN) for accuracy. Baseline data items which will be extracted will include:
  - First author name, year and journal of publication
  - Language of publication
  - Study design: human/cadaveric/animal/experimental; whether comparative or non-comparative; whether prospective or retrospective (for human studies).
  - Number of patients/experiments
  - Orthopedic surgical procedure investigated
  - Orthopedic power tools/ instruments investigated
f. *Outcome Measures* Owing to the broad nature of the research question, we acknowledge that it is difficult to precisely define all the possible outcome measures. However, the following important outcome measures will be included, others may be added as the review evidence is gathered :
  - Total and/or viable particle counts
  - Aerosol particle size
  - Aerosol particulate concentration
  - Microbiological air contamination
  - Aerosol scatter characteristics
  - Contamination of OR personnel by aerosols
  - Presence of blood in aerosols
g. Data Analysis and Synthesis Both qualitative and quantitative analyses will be performed. For qualitative analysis, appropriate tables and data visualization diagrams will be constructed. Baseline data items as well as all the pre-specified outcome measures will be reported. Wherever appropriate, meta-analysis using a *random-effects* model will be used to calculate pooled effect-size estimates of outcome variables (for e.g. total particle count, particle density etc.); results will be reported as means with 95% confidence intervals. Forest plots will be constructed to visualize the results. Statistical heterogeneity will be evaluated by the *I*^2^ test. If high statistical heterogeneity is identified (I^2^ > 75%), leave-one-out sensitivity analysis will be performed to identify the effect of each included study on the overall effect estimate. No subgroup analysis has been planned *a-priori*; however, it may be undertaken depending on the available evidence. Analysis will be performed by the *Open Meta Analyst Software*.
h. *Assessment of Risk of Bias* The quality of the studies included will be evaluated by the Risk of Bias in Studies estimating Prevalence of Exposure to Occupational risk factors (RoB-SPEO) tool (6). The studies will be individually rated as *‘low, probably low, probably high, high or no information’* based on bias assessment in eight separate domains. Although this tool is relatively new and has limited peer validation, it is well-suited for the purposes of this systematic review, and has been shown to have good inter-observer agreement.
i. Assessment of Strength of Evidence The overall strength of evidence will be assessed by the GRADE approach.

## Data Availability

Since this is a protocol for systematic review, it does not contain data. However, the search strategy has been presented in Table 1.

## APPENDIX 1 PRISMA-P (Preferred Reporting Items for Systematic review and Meta-Analysis Protocols) 2015 checklist: recommended items to address in a systematic review protocol*

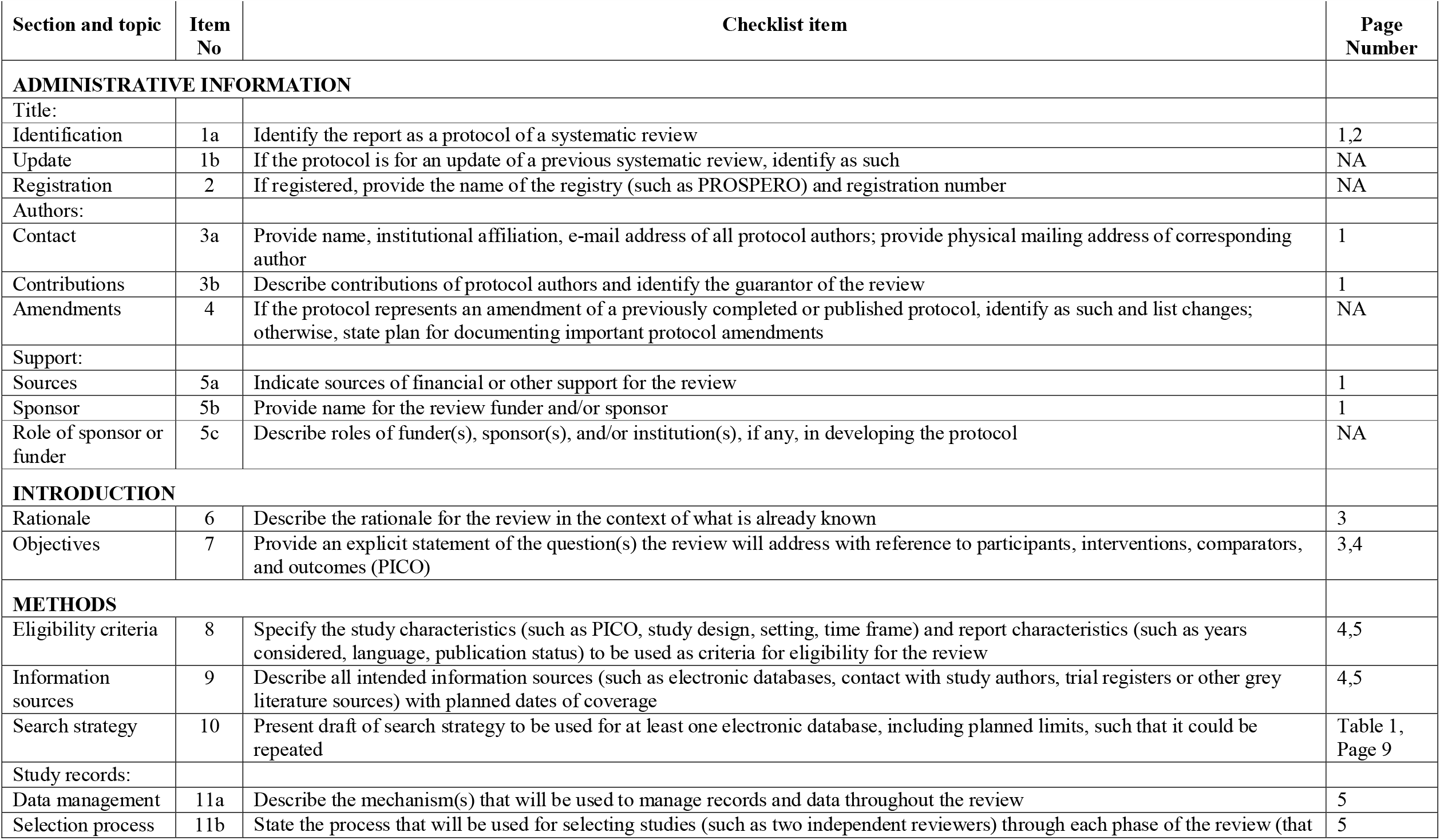
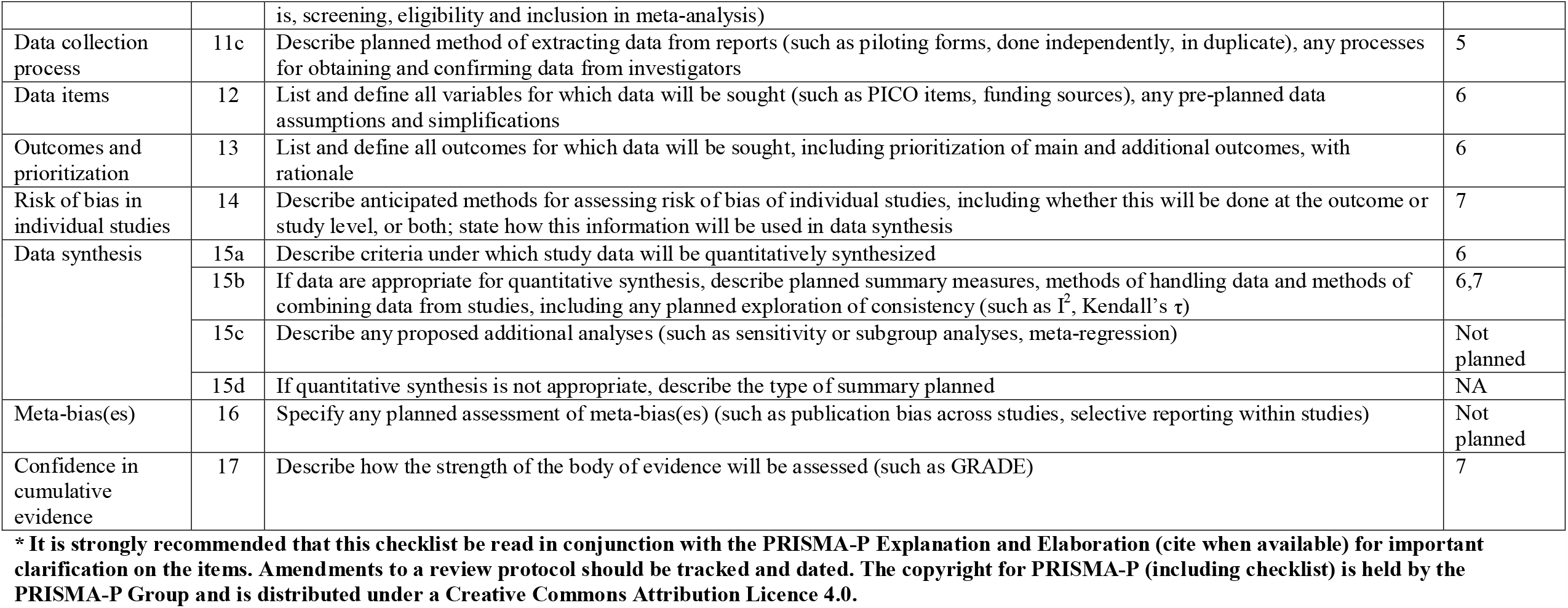

| Section and topic | Item No | Checklist item | Page Number |
| --- | --- | --- | --- |
| **ADMINISTRATIVE INFORMATION** |  |  |  |
| Title: |  |  |  |
| Identification | 1a | Identify the report as a protocol of a systematic review | 1,2 |
| Update | 1b | If the protocol is for an update of a previous systematic review, identify as such | NA |
| Registration | 2 | If registered, provide the name of the registry (such as PROSPERO) and registration number | NA |
| Authors: |  |  |  |
| Contact | 3a | Provide name, institutional affiliation, e-mail address of all protocol authors; provide physical mailing address of corresponding author | 1 |
| Contributions | 3b | Describe contributions of protocol authors and identify the guarantor of the review | 1 |
| Amendments | 4 | If the protocol represents an amendment of a previously completed or published protocol, identify as such and list changes; otherwise, state plan for documenting important protocol amendments | NA |
| Support: |  |  |  |
| Sources | 5a | Indicate sources of financial or other support for the review | 1 |
| Sponsor | 5b | Provide name for the review funder and/or sponsor | 1 |
| Role of sponsor or funder | 5c | Describe roles of funder(s), sponsor(s), and/or institution(s), if any, in developing the protocol | NA |
| **INTRODUCTION** |  |  |  |
| Rationale | 6 | Describe the rationale for the review in the context of what is already known | 3 |
| Objectives | 7 | Provide an explicit statement of the question(s) the review will address with reference to participants, interventions, comparators, and outcomes (PICO) | 3,4 |
| **METHODS** |  |  |  |
| Eligibility criteria | 8 | Specify the study characteristics (such as PICO, study design, setting, time frame) and report characteristics (such as years considered, language, publication status) to be used as criteria for eligibility for the review | 4,5 |
| Information sources | 9 | Describe all intended information sources (such as electronic databases, contact with study authors, trial registers or other grey literature sources) with planned dates of coverage | 4,5 |
| Search strategy | 10 | Present draft of search strategy to be used for at least one electronic database, including planned limits, such that it could be repeated | Table 1, Page 9 |
| Study records: |  |  |  |
| Data management | 11a | Describe the mechanism(s) that will be used to manage records and data throughout the review | 5 |
| Selection process | 11b | State the process that will be used for selecting studies (such as two independent reviewers) through each phase of the review (that is, screening, eligibility and inclusion in meta-analysis) | 5 |
| Data collection process | 11c | Describe planned method of extracting data from reports (such as piloting forms, done independently, in duplicate), any processes for obtaining and confirming data from investigators | 5 |
| Data items | 12 | List and define all variables for which data will be sought (such as PICO items, funding sources), any pre-planned data assumptions and simplifications | 6 |
| Outcomes and prioritization | 13 | List and define all outcomes for which data will be sought, including prioritization of main and additional outcomes, with rationale | 6 |
| Risk of bias in individual studies | 14 | Describe anticipated methods for assessing risk of bias of individual studies, including whether this will be done at the outcome or study level, or both; state how this information will be used in data synthesis | 7 |
| Data synthesis | 15a | Describe criteria under which study data will be quantitatively synthesized | 6 |
| | 15b | If data are appropriate for quantitative synthesis, describe planned summary measures, methods of handling data and methods of combining data from studies, including any planned exploration of consistency (such as $I^2$, Kendall's $\tau$) | 6,7 |
|  | 15c | Describe any proposed additional analyses (such as sensitivity or subgroup analyses, meta-regression) | Not planned |
|  | 15d | If quantitative synthesis is not appropriate, describe the type of summary planned | NA |
| Meta-bias(es) | 16 | Specify any planned assessment of meta-bias(es) (such as publication bias across studies, selective reporting within studies) | Not planned |
| Confidence in cumulative evidence | 17 | Describe how the strength of the body of evidence will be assessed (such as GRADE) | 7 |

**\* It is strongly recommended that this checklist be read in conjunction with the PRISMA-P Explanation and Elaboration (cite when available) for important clarification on the items. Amendments to a review protocol should be tracked and dated. The copyright for PRISMA-P (including checklist) is held by the PRISMA-P Group and is distributed under a Creative Commons Attribution Licence 4.0.**

*From: Shamseer L, Moher D, Clarke M, Ghersi D, Liberati A, Petticrew M, Shekelle P, Stewart L, PRISMA-P Group. Preferred reporting items for systematic review and meta-analysis protocols (PRISMA-P) 2015: elaboration and explanation. BMJ. 2015 Jan 2;349(jan02 1):g7647*.

## Abbreviations

AGP: Aerosol Generating Procedure(s)
OR: Operating Room
OSP: Orthopedic Surgical Procedure(s)
PRISMA: Preferred Reporting Items for Systematic review and Meta-Analysis
PRISMA-P: Preferred Reporting Items for Systematic review and Meta-Analysis - Protocols
RoB-SPEO: Risk of Bias in Studies Estimating Prevalence of Exposure to Occupational Risk Factors
SARS-CoV-2: Severe Acute Respiratory Syndrome Coronavirus 2
COVID-19: Coronavirus Disease

